# Hacking Acute Care: A Qualitative Study on the Healthcare Impacts of Ransomware Attacks Against Hospitals

**DOI:** 10.1101/2023.02.13.23285854

**Authors:** Liselotte S. van Boven, Renske W.J. Kusters, Derrick Tin, Frits H. M. van Osch, Harald De Cauwer, Linsay Ketelings, Madhura Rao, Christian Dameff, Dennis G. Barten

## Abstract

**Background and objectives:** Cyberattacks against healthcare institutions are an increasing threat and have the potential to impair health outcomes. Current research is limited and focuses mainly on the technical consequences, whereas little is known about the healthcare staff experiences and the impact on emergency care, both during the incident and in the recovery phase. This study aims to explore the impact of a sample of large ransomware attacks against hospitals between 2017 and 2022 on acute care delivery and patient care during the recovery phase.

**Methods:** This interview-based qualitative study assessed the experiences of emergency healthcare professionals and Information and Communication Technology (ICT) staff and investigated the challenges faced when struck by a major hospital ransomware attack. The semi-structured interview guideline was based on current literature and cybersecurity expert consultation. Transcripts were anonymized and information tracing back to participants and/or their organizations was removed for privacy purposes.

**Results:** Nine participants were interviewed, including emergency healthcare providers and ICT-focused staff. Five themes were constructed from the data: impact and challenges regarding patient care continuity, challenges during the recovery process, personal impact on healthcare staff, preparedness, and lessons identified and future recommendations.

**Conclusions:** According to the participants of this qualitative study, ransomware attacks have a significant impact on emergency department (ED) workflow, acute patient care and the personal wellbeing of healthcare providers. Preparedness for such incidents is often limited and many challenges are encountered during the acute and recovery phase of the attack. Proactive preparedness efforts are essential to improve contingency planning and to develop response strategies for hospital ransomware attacks.

## Introduction

Along with the digital revolution in healthcare, cybercrime is a rapidly emerging threat to healthcare security. Healthcare institutions, and hospitals in particular, often lag behind with data protection because cybersecurity investments lack priority, ^1^ making them soft targets for digital crime.

Recently, ransomware attacks in the European healthcare sector have reached an unprecedented high. ^2^ Ransomware is a type of malware that intentionally inserts software into a system for harmful purposes. ^3^ It specifically encrypts data, rendering data on infected computerized systems inaccessible until a ransom payment is made to the perpetrators, ^4^ typically in cryptocurrency. Systems are often taken offline by the targeted organization as a protective strategy to prevent further spread of malware through unaffected systems. Cyberattacks targeting hospitals are so-called “threat-to-life crimes”, ^5^ because disruption of patient care potentially has severe consequences on patient well-being and may lead to impaired health outcomes and increased mortality. ^6 7 8 9^ Healthcare workers previously reported longer emergency department (ED) stay, delayed testing and treatment, increased necessity for patient transfers and higher complication rates. ^10^ Since hospitals are highly dependent on continuity of care, stakes are high and cybercriminals know healthcare institutions will go to great lengths to ensure the restoration of daily practice. ^11 12^

The coronavirus disease 2019 (COVID-19) pandemic may have acted as a smokescreen for cybercrime, keeping hospitals’ attention focused on the pandemic. The World Health Organization (WHO) reported a five-fold increase in cyberattacks against healthcare institutions during the COVID-19 pandemic. ^13^ The pandemic and the associated stress possibly made individual healthcare workers more vulnerable to scams. More healthcare staff worked from home using less secure networks or devices, using online platforms for digital meetings that are prone to hacking. ^14 15^ In addition, hackers used cyberscams to exploit people’s fear by spreading phishing emails with fake COVID-19 information updates. ^16^ Previous studies have shown that poor user awareness (among healthcare staff) is one of the most important risks to healthcare organizations. Personalized education campaigns may therefore be the most efficient protection strategy against phishing attacks. ^16^

Existing evidence on the healthcare impacts of ransomware attacks is limited. The majority of studies focused on cybercrime trends over the past years, ^17^ the financial impact of cybercrime, ^18 19^ Information and Communication Technology (ICT) related aspects ^1^ or specific cyber incidents. ^20 21^ However, little is known about the continuity of patient care and the recovery of acute care delivery during and after a ransomware attack. ^22^ The current study aims to characterize the acute care impact of several large ransomware attacks against hospitals in recent years. This may provide insights to improve hospital preparedness for ransomware attacks.

## Methods

The study is reported using the Consolidated criteria for Reporting Qualitative research (COREQ). ^23^

### Study Design

A qualitative study was performed using semi-structured interviews with healthcare professionals and ICT staff to explore their experiences and roles as well as the challenges faced during and after ransomware attacks. The phenomenological approach of the interviews allowed an in-depth investigation of participants’ lived experiences.

### Incident Inclusion and Definitions

The authors constructed a list of large ransomware attacks against hospitals in Europe and the United States between 2017 and 2022 based on a review of literature and news articles on Google News, Google Scholar, PubMed and LexisNexis. Additionally, the Dutch Computer Emergency Response Team (Z-CERT) provided online sources listing healthcare organizations targeted by ransomware attacks. Incidents in this list that met the inclusion criteria were selected and the associated healthcare organizations were subsequently contacted. The list of incidents was reviewed by a multidisciplinary panel of 5 experts in the fields of cybersecurity, emergency medicine, disaster medicine and counter-terrorism medicine. The composition of this panel was based upon relevant peer-reviewed publications and expert opinion (advice from co-authors and Z-CERT). For the purposes of this study, large ransomware attacks were defined as ‘attacks with a serious disruption of acute care’, including in-patient transfers, disruption of acute care services and loss of access to Electronic Health Records (EHR). Incidents with minimal or no disruption to care delivery are often not reported in literature or by media platforms and were therefore not included.

### Participant Recruitment and Selection

Target participants included crisis coordinators, acute care department staff and (ICT) managers involved in these incidents. Purposive sampling and snowball sampling techniques were used to reach the most suitable participants. ^24^ These sampling methods are the most efficient way to gather information of interest as the study field has a limited number of individuals who can serve as primary data sources. Targeted organizations were either contacted using a general email address or, if specific contact information could be obtained, by directly contacting the ED and/or medical informatics department. In some cases, one or more or the authors had contacts in a targeted organization and approached the participants. Interviews were carried out until data saturation was achieved.

### Data collection

Data collection consisted of semi-structured interviews. The interview guideline (Appendix 1) was designed for this study by the research team, based on literature research and cybersecurity expert consultation. Literature review provided insight into the aspect of clinical work most immediately affected by cyberattacks and provided a useful basis for the interview questions. The guideline was not piloted before use in the study and remained the same throughout the interviews. Interviews were conducted in English and Dutch by a single interviewer (LvB, female, Master of Medicine) via video calls using Zoom (© Zoom Video Communications, Inc., Version 5.9.6). The interviews were performed in April and May 2022 after obtaining informed consent. Interviews consisted of open-ended prompt questions regarding the themes as presented in the interview guideline (Appendix 1). Probing questions provided a more in-depth discussion. No repeat interviews were conducted. Interviews were recorded using voice-recording software (©Apple Voice Memos, Apple Inc., Version 2.3), no notes were taken during the interviews. To ensure data privacy and participant anonymity, audio recordings and interview transcripts were only available to the executive researcher (LvB) and stored on a password-protected computer. Transcripts were anonymized and any traceable information to the participant or the associated organization was omitted. Considering the sensitive nature of ransomware attacks on hospitals, technical details of the attacks were asked in general and categorical terms and the focus was primarily on the impact on patient care. No prior relationship existed between the interviewer and the participants. Participants were informed of the general purpose and background of the study.

### Data analysis

The voice-recorded interviews were transcribed *verbatim* in Microsoft Word (Microsoft ® for Mac, Version 16.60), anonymized and returned to the participants for a factual check before data analysis. The transcripts were imported into the Atlas.ti software program (Atlas.ti Scientific Software Development GmbH, Version 22.0.1). All transcripts were coded using in vivo coding by one author (LvB) as participants’ own words were used as code titles. ^25^ The final stage of data analysis included the identification of suitable quotes which were validated with participants before use in the final article. Participants were able to provide feedback and ensure a correct interpretation of their experiences and opinions was obtained.

### Ethical Considerations

The study was approved by the Medical Ethical Committee (METC) of the Maastricht University Medical Center (MUMC) (METC 2022-3106) and the Maastricht University FHML Research Ethics Committee (FHML-REC/2022/015).

## Results

### Characteristics of study participants

Twenty-five hospital institutions were invited to participate. Of these, 18 did not respond and three did not wish to participate because of a lack of time (n=1), ongoing police investigations into the incident (n=1) or concerns regarding the sensitivity of the subject and data privacy (n=1) despite extensive measures taken by the researchers to ensure data anonymity. In total, nine individuals were interviewed from four different organizations that sustained a ransomware attack between 2018 and 2022. Participant and incident demographics are outlined in Table 1 and Table 2. Interviews had a mean duration of 52 minutes (range 27 to 83 minutes). The coding process was repeated twice to identify remaining codes. Similar codes within the 89 assigned codes were linked, resulting in 14 categories. These categories were organized into five themes (Table 3). Data saturation was determined during the coding process and was reached after six interviews.

**Table 1:** Participant demographics

| **Participant** | **Gender<sup>1</sup>** | **Age** | **Role during ransomware incident** | **Years of experience<sup>2</sup>** | **Ransomware incident number<sup>3</sup>** |
| --- | --- | --- | --- | --- | --- |
| P1 | M | 36-40 | Emergency Physician | 14 | R1 |
| P2 | F | 55-60 | Manager / hospital administration | >20 | R1 |
| P3 | F | 30-35 | Emergency Physician | 6 | R2 |
| P4 | M | 35-40 | Manager / hospital administration | 18 | R3 |
| P5 | M | 45-50 | Emergency Physician | >20 | R4 |
| P6 | M | 50-55 | Manager / hospital administration | 27 | R1 |
| P7 | M | 30-35 | Emergency Physician | 12 | R4 |
| P8 | F | 40-45 | Emergency Physician | 10 | R3 |
| P9 | M | 60-65 | Manager / hospital administration | 38 | R3 |

<sup>1</sup>M = male, F = female
<sup>2</sup>Note: this is an approximation.
<sup>3</sup>Ransomware incident number corresponds to the number assigned to each incident, as shown in Table 2
Note: none of the participants experienced a previous malware or ransomware incident.

**Table 2:** Hospital and Ransomware incident demographics

| **Ransomware incident** | **R1** | **R2** | **R3** | **R4** |
| --- | --- | --- | --- | --- |
| **Type of hospital<sup>1</sup>** | Secondary | Tertiary | Tertiary | Tertiary |
| **Number of inpatient beds<sup>2</sup>** | 300-400 | 300-400 | 300-400 | 200-300 |
| **Catchment area<sup>2</sup>** | >100,000 | Unknown, ED serves ±200 patients per day | >100,000 | >100,000 |
| **Trauma level** | 2 | 1 | 1 | 2 |
| **Other tertiary functions<sup>3</sup>** | ICU, PCI center | ICU, PCI center, stroke center | ICU, PCI center, NICU, PICU, CCU | ICU, PCI center (diagnostic only), NICU, stroke center |
| **Duration of incident<sup>4</sup>** | >6 weeks<sup>a</sup> | 2-3 weeks | 3-4 weeks | 2-3 weeks |
| **Loss of access to computers** | Yes | Yes | Yes | No<sup>b</sup> |
| **Loss of access to EHR** | Yes | Yes | Yes, ICT staff shut down EHR system for safety measures | Yes |
| **Loss of access to internal telephone system** | Yes | Yes | Yes | No |
| **Loss of internet access** | Yes | Yes | Yes, hospital ICT staff shut down the internet for safety measures | Yes |
| **Loss of access to hospital digital medication system** | No, hospital did not employ digital medication system | Yes | Yes | No, hospital did not employ digital medication system |
| **Loss of access to digital radiology system** | Yes | Yes | Yes | Yes |
| **Loss of access to digital laboratory system** | Yes | Yes | Yes | Yes |
| **Affected department(s)** | All | All | All | All |
| **Cancellation of outpatient appointments** | Yes | Unknown | Yes | Unknown |
| **Hospital diversion** | No | Yes, ambulance diversion during first 24 hours | No | No |
| **ED closure** | No | No | No | No |
| **OR closure** | Partial; planned operations were cancelled but emergency operations were carried out | Partial; some planned operations were cancelled but emergency operations were carried out | Partial; planned operations were cancelled | No |
| **Data leak<sup>5</sup>** | No | No | No | No |
| **Patient harm<sup>5</sup>** | No | No | No | No |

<sup>1</sup>In order to maintain incident anonymity, European and U.S. care systems were merged for the purpose of this article. European peripheral hospitals and teaching hospitals fall under 'secondary', European academic hospitals fall under 'tertiary'.
<sup>2</sup>Expressed categorically to maintain incident anonymity.
<sup>3</sup>ICU = intensive care unit, PCI center = percutaneous coronary intervention center, CCU = cardiac care unit, NICU = neonatal intensive care unit, PICU = pediatric intensive care unit
<sup>4</sup>Expressed categorically to maintain incident anonymity. Defined as period during which hospital computer systems were not functioning as usual.
<sup>5</sup>As reported by participants and/or news articles covering the incident.
<sup>a</sup>Incident R1 had a duration of several months. This was largely due to a criminal investigation launched by authorities regarding the ransomware attack. Access of hospital IT staff to the system was denied, as this may have interfered with the ongoing investigation. However, this also meant that IT recovery could not take place and the downtime was significantly extended.
<sup>b</sup>Incident R4 involved a loss of internal internet access. Therefore, computers were available, but any application or functionality requiring internet access was unavailable.

**Table 3:** Themes and categories identified during data analysis

| Theme | Category |
| --- | --- |
| Patient care continuity | Lack of electronics<br>Communication<br>Diagnostics: lab and radiology<br>Patient safety<br>Inadequate emergency plans<br>Adaptations |
| Challenges during recovery process | Challenging ICT recovery process<br>Difficult return to digital system |
| Personal impact on healthcare staff | Emotional impact |
| Preparedness | Preparedness for cyberattack |
| Lessons identified and future recommendations | Improved communication<br>Improved ICT security<br>Improved contingency plans<br>Changes made since incident |

### Main results

#### Patient care continuity

##### Lack of electronics

The most notable impact of the ransomware attacks was the loss of technology availability, as a direct result of the attack or as a preventive measure taken by ICT staff. Three out of four incidents involved complete computer downtime. In most cases, email and internal hospital telephone systems were unavailable. Emergency care protocols were unavailable during the attacks and had to be retrieved from the internet or borrowed from other hospitals. Hospital staff often felt lost without the availability of computers and described it to be crippling: “*hospitals have become so digital that you can’t really work without these systems and that’s a scary thought*”. Similarly, someone stated that “*the expectations for the standard of care here are dependent on real-time electronic connection*”.

All hospitals experienced a switch to a paper-based charting system. Some hospitals had paper charting forms available but staff was often not instructed where to find them: “*it was a scramble to find stuff*”. Paper charting forms were very basic and not designed for ‘modern’ users accustomed to electronic charting methods. Most participants regarded paper charting as inefficient and reported decreased work productivity. Whereas older staff were often familiar with paper charting, young professionals had never practiced medicine in a paper environment and frequently needed assistance from senior staff. The need for paper charting also impacted providers’ ability to keep track of patients: “*you don’t really know who’s in the waiting room*”. All hospitals replaced the digital ED tracking board with a whiteboard. However, constant whiteboard edits were confusing and ED staff indicated difficulties in tracking and reporting patient status.

##### Communication

The lack of communication tools such as internal telephone systems and email necessitated the use of alternative communication methods such as fax machines, the hospital’s pneumatic tube system, personal smartphones, walkie-talkies and pagers. A participant mentioned, “*I still think some of the biggest hurdles and difficulties were interacting with other divisions and departments; lab, radiology and follow-up*”. Internal hospital communication surrounding the incident was often limited. Several participants found out about the incident via local news and said, “*administration just wasn’t telling us what was going on*”. Updates often took place via word of mouth and one hospital employed a mass-messaging system to employees’ personal phones.

##### Diagnostics: lab and radiology

Difficulties ordering and obtaining diagnostics were common. Carrying forms back and forth and reviewing radiology images in person in the radiology department was time-consuming. Most hospitals redeployed non-clinical personnel as ‘runners’ for those who delivered care. “*The attending physician had to read through all the results and make sure there wasn’t anything serious in there, and sign them all to say that they were received, and then put them into a pile and the residents would get their labs out of the pile*”. Diagnostic orders were limited to emergent matters: “*we modified our ordering patterns within the ED, and limited it to emergent things in order to offload lab and radiology*”.

##### Patient safety

Multiple participants described that ED patient interaction was not altered, but concerns arose about the inaccessibility of patient history and medication lists. Participants felt it hampered care provision because it was hard to verify patients’ personal accounts. In addition, the ransomware attacks “*caused massive delays*” in patient care and providers worried that these delays led to worse outcomes. Similarly, someone stated, “*it clearly caused harm, just in terms of delay of patient care and not knowing about their follow-up*”. None of the ransomware attacks were directly linked to a case of patient harm, but staff deemed it a high-risk situation. One physician mentioned the concern of important diagnostic findings going missing or potentially overlooking a patient with a serious and acute condition in the waiting room due to crowding. Patients may have been lost to follow-up and did not receive the appropriate care as a result. Lastly, digital handover of patients was impossible: “*especially in the extensiveness, the tidiness, the legibility of patient handover, there was a lot of information loss there*”. Most hospitals continued the provision of acute care services during the ransomware event. In one incident, the ED went on diversion during the first 24 hours of the incident.

##### Inadequate emergency plans

A challenge encountered by healthcare providers and ICT staff was the limited applicability of the downtime plans to prolonged downtime: “*we learned that the realities of a cyber event stress some of the expectations and assumptions within downtime procedures*”. Downtime plans often did not account for a disaster in which triage needs to be based upon the availability of technological applications. Paper plans were often focused on EHR downtime but not on the downtime of all ICT systems. Although some incidents had reverted to downtime procedures in the past, little has been done to optimize these procedures.

##### Adaptations

Some hospitals called extra staff members to work, or redeployed staff to more vital areas: “*the hospital provided additional non-medical staff to perform tasks that really didn’t need to be done by our clinical staff*”. Despite the many challenges the ransomware attacks posed, most teams gradually improved and developed more streamlined care processes. A commonly shared opinion was the adaptability of ED staff to new and unknown situations: “*we’re used to being thrown into unknown circumstances. It’s a little bit of what our profession is based on*”.

#### Challenges during recovery process

##### Difficulties in ICT recovery process

Three participants were involved in the ICT aspects of the attacks. Establishing priority in recovering the system’s applications was a major challenge during the recovery phase: “*when you’re in it, you realize there’s really only one prioritization question, and that’s ‘how vital is this to patient care?*’”. Another challenge was that the recovery of one application often depended on the recovery of several supporting applications. In one incident, a forensic investigation withheld the ICT department from access to the systems and carrying out the planned disaster recovery. It took months for authorities to clear the systems: “*my systems were being held hostage, and not only by the ransomware*”.

##### Difficulties with return to digital system

The return to a digital system after restoring computer and EHR access was associated with several challenges. As the computers were gradually restored, determining staff priority regarding EHR access was difficult: “*different departments were trying to wrestle for who got priority*”. Information loss was a commonly discussed issue: some of the paper charts were lost, paper charting was incomplete or illegible and several digital files could not be recovered after the incident.

Furthermore, the back-entering of data was a labor-intensive and time-consuming task. Many files were scanned into patients’ digital files to save time and only the most relevant things were back-entered manually. One hospital paid personnel to work extra hours and enter all diagnostic and medication orders manually before the relaunch of the digital system.

#### Personal impact on healthcare staff

Many participants experienced the ransomware incident as emotionally impactful and described it as a “*miserable occurrence*” and “*emotionally, a rollercoaster*”. Staff was fearful of patient harm and ICT staff felt their inability to recover certain applications could impact patient lives: “*I felt responsible in some ways because I work for IT and we allowed this to happen*”. Similarly, someone stated: “*if we don’t recover this application as soon as possible, it could cost human lives*”. Providers were also fearful that patient and personal data would end up in the wrong hands and/or be made public. The increased workload led to increased stress: “*I worked twenty hours a day, seven days a week for five weeks*”, “*you don’t sleep, you’re working 24/7*”. In most cases, COVID-19 added stress: *“everybody was already fried, exhausted, at their wits’ end, burnt out*”. Furthermore, participants said hospital staff was embarrassed that they couldn’t care for patients properly due to the lack of resources. An ICT manager felt derided by other organizations: “*people are going to ridicule you; shaming and blaming*”. Contrastingly, participants also indicated some positive outcomes, such as an increased sense of “*comradery*” within the institution: “*the organization has probably never been as aligned, or as in-sync, and operating as one as during that cyberattack*”. One participant said: “*I love acute things, so it was a mighty experience*”.

#### Preparedness

Most participants indicated that preparedness was limited and they “*were coming in blind*”. A participant indicated that the organization was not well-prepared for weeks of outage. They said: *“we did technically have a downtime plan in place but not something that would have been usable for multiple days to weeks on end*”. One ED physician deemed the hospital’s ICT department “*incompetent*”. Contrarily, one participant felt that her organization was above average prepared due to their focus on disaster medicine and some stated that previous short-term downtimes prepared them for a ransomware attack to some extent. One participant with an ICT background stated that the ICT planning and preparation for a cyberattack was adequate.

#### Lessons identified and future recommendations

##### Improved communication

Multidisciplinary communication proved difficult: “*there needs to be seamless communication between cyber-event planning and emergency management planning*”. Participants encouraged transparency to hospital staff and patients to avoid uncertainty and frustration. The use of walkie-talkies, pagers, personal smartphones, fax machines and mass-messaging systems proved useful.

##### Improved ICT security

All participants voiced a need for improved ICT security with regular system updates and balanced investments into hospital ICT systems and care-related issues. Early detection of suspicious activity would be very useful in prevention or early response measures. Another important lesson was the segmentation of a system’s critical servers, or “*crown jewels*”, meaning the servers are placed behind an extra ‘safety wall’ and are not easily accessible. Furthermore, the prioritization of applications could be improved. Finally, creating regular backups of patient data would increase preparedness.

##### Improved contingency plans

A crucial lesson was the need for improved contingency plans. This involved revamping of downtime plans, documenting the new workflow and adjusting downtime forms. Practicing downtime planning and performing table-top scenario training was expected to drastically improve preparedness for cyberattacks: “*the exercise of planning, of discussing it with each other, of scenarios and finding solutions to them is the most important thing, not the plan itself*”. Several recommendations were made to ensure patient care continuity. One recommendation was the formation of ‘runners plans’; establishing which personnel can be diverted to support the most essential functions. In addition, pre-establishing relationships with institutions that can aid in the recovery process is beneficial. Putting the ED on temporary diversion may provide time for the establishment of an effective workflow and to avoid ED overcrowding. Storing paper charting forms may aid in a quick and efficient switch to paper charting. One participant suggested a bank of laptops working on an independent server that serves as a mobile cyber response unit.

##### Changes made since incident

Most hospitals made changes since the ransomware attack in accordance with the aforementioned lessons identified. Hospitals set up more secure log-in procedures, applied extensive limitations to remote system access, encrypted classified information and, in some cases, ICT staff now carry out regular ‘penetration testing’ of the system to identify vulnerabilities. A vital change was increasing awareness and training of hospital staff by sending out fake phishing emails, providing warnings about non-secure websites, requiring a regular change of password, yearly cybersecurity training and e-learning courses. One participant organized a cyber awareness event, “*about ransomware, phishing, sextortion, cyberjacking*”. Scenario-based training was organized, such as regular fail-overs of the EHR to test ICT staff response to system downtime. Despite the ransomware attack, some organizations made no concrete improvements.

## Discussion

This study assessed the impact of ransomware attacks against hospitals on acute care delivery. Although most hospitals in this study continued acute care, participants reported a major and immediate impact on acute care services and personal wellbeing. Participants were concerned about patient safety and most hospitals were ill-prepared for prolonged downtime procedures.

The extensive personal impact of ransomware attacks on healthcare workers in this study is consistent with the findings of Carton and co-workers, who assessed the ransomware attack on the Health Service Executive (HSE) Ireland. ^26^ Participants mentioned emotions such as fear, stress, sadness, anger and disgust. Poor mental health amongst healthcare providers is significantly associated with decreased patient safety and more self-reported errors. ^27^ Therefore, attention to staff wellbeing is crucial in times of crisis. A previous study on employee vulnerability to phishing scams demonstrated a positive association between staff workload and the likelihood of clicking on phishing emails. ^22^ As a result, cyberattacks may lead to decreased mental well-being of hospital staff, therefore contributing to cyberattack vulnerability and possibly increasing patient safety risks.

Any disruption of acute care services may impact patient care and safety ^28^. Researchers recently found increased mortality rates in hospitals targeted by cyberattacks, further stressing the gravity of the issue. ^9^ All participants in this study expressed concerns and suspicions regarding decreased patient safety due to the incident, although no root-cause analyses were performed. A study that assessed safety event reports already demonstrated the importance of health ICT to patient safety and quality of care. ^29^ A large number of ICT failure incidents led to patient harm and 75% of incidents were deemed preventable. Consistent with these findings, participants in this study stressed the importance of ICT investment and cyber-specific contingency plans to prevent cyberattacks. Acute care delay was an important factor regarding patient safety concerns in this study. Previous studies showed that delayed ED care impacts patient safety, mainly due to longer time to triage and time to care provision. ^28^ Improved preparedness may lead to better downtime workflow and fewer delays in care, thereby limiting the impact of a cyberattack on patient safety.

A consensus between all participants existed about the limited hospital preparedness for a cyberattack. A preparedness survey of healthcare employees by Branch and co-workers found that emergency plans are often present but not specific for cyber events. ^30^ One-third of responders rated the organization’s overall preparedness as limited. Contrary to this study, ICT personnel generally had more confidence in organization preparedness than healthcare staff. ^30^ A possible explanation might be that the responders in the survey did not experience a cyberattack. ^30^ Consistent with the literature, this study demonstrates limited cybersecurity awareness and the importance of staff awareness and response training. ^17 26 27 31^ Active scenario-based staff training and system testing may improve hospital preparedness for future cyberattacks. ^32^

This study builds upon existing evidence by providing recommendations specific to patient care. Key aspects are teaching staff paper-based charting skills and having paper copies of charts and diagnostic order forms at hand. Having protocols available when the hospital system is down may ameliorate patient care, and temporary ED diversion during the first hours of the cyberattack may reduce pressure on acute care services and provide time to transition into downtime workflow. Reverse triage, in which patients already present at the facility are transferred to uninfected hospitals, may alleviate pressure on the infected location. Above all, transparency to hospital staff as well as to patients and external partners is pivotal in avoiding uncertainty and mitigating negative personal impacts.

## Strengths and Limitations

There are several limitations associated with this study. Although most healthcare organizations recognized the priority of the topic, the willingness to contribute was limited. This could be explained by concerns regarding the sensitivity of the topic, despite meticulous efforts to ensure data anonymity. Reasons for participation denial may include security reasons, liability concerns, and sense of failure. Nevertheless, the number of participants was sufficient to achieve data saturation. It should be noted that incidents were only included if they were associated with a significant disruption of hospital care. Statements about healthcare impact should therefore be read with caution, as the argument may become circular. However, the inclusion of major ransomware attacks was a deliberate strategy, as the authors assumed that minor incidents were less often reported in news articles.

Another limitation is potential researcher bias due to the subjective nature of the study’s qualitative design. In addition, interviews and coding were performed by a single researcher. The main researcher’s (LvB) medical background introduces potential researcher bias, including data collection- and analysis bias. These effects were minimized by the use of a standardized interview guide and the employment of in vivo coding. Selection bias is an issue inherent to purposive sampling. However, this method was intentionally chosen to obtain participants with the potential to provide information relevant to the research question. ^33^ Additionally, recall bias may have occurred as the incidents took place in the past 5 years. Lastly, the relatively small sample of incidents (n=4) may contribute to the limited impacts found on acute care in terms of ED closure or ambulance diversion. However, it is not necessarily the sample size that determines the quality of qualitative research, but rather the depth accomplished within the research. ^34^ In terms of data adequacy, data richness is an equally important marker as sample size. The detailed study findings allowed the researchers to interpret the meaning and context of the identified themes. ^35 36^ This study also has several strengths. First, this research assessed the healthcare impact of ransomware attacks, which has not been widely investigated despite the daily threats and attack risks to modern healthcare. Furthermore, healthcare staff with varying backgrounds and positions were interviewed, providing a multidisciplinary point of view. The inclusion of incidents across Europe and the United States strengthened external validity. Internal validity was improved by using a standardized interview guide, member checking during the interviews and participant validation of interview transcripts.

## Conclusion

According to the participants of this qualitative study, ransomware attacks can have a significant impact on emergency department (ED) workflow, acute patient care and the personal wellbeing of healthcare providers. Preparedness for such incidents is often limited and many challenges are encountered during the acute and recovery phase of the attack. Proactive preparedness efforts are essential to improve contingency planning and to develop response strategies for hospital ransomware attacks.

## Data Availability

All data produced in the present work are contained in the manuscript

**Appendix 1:**
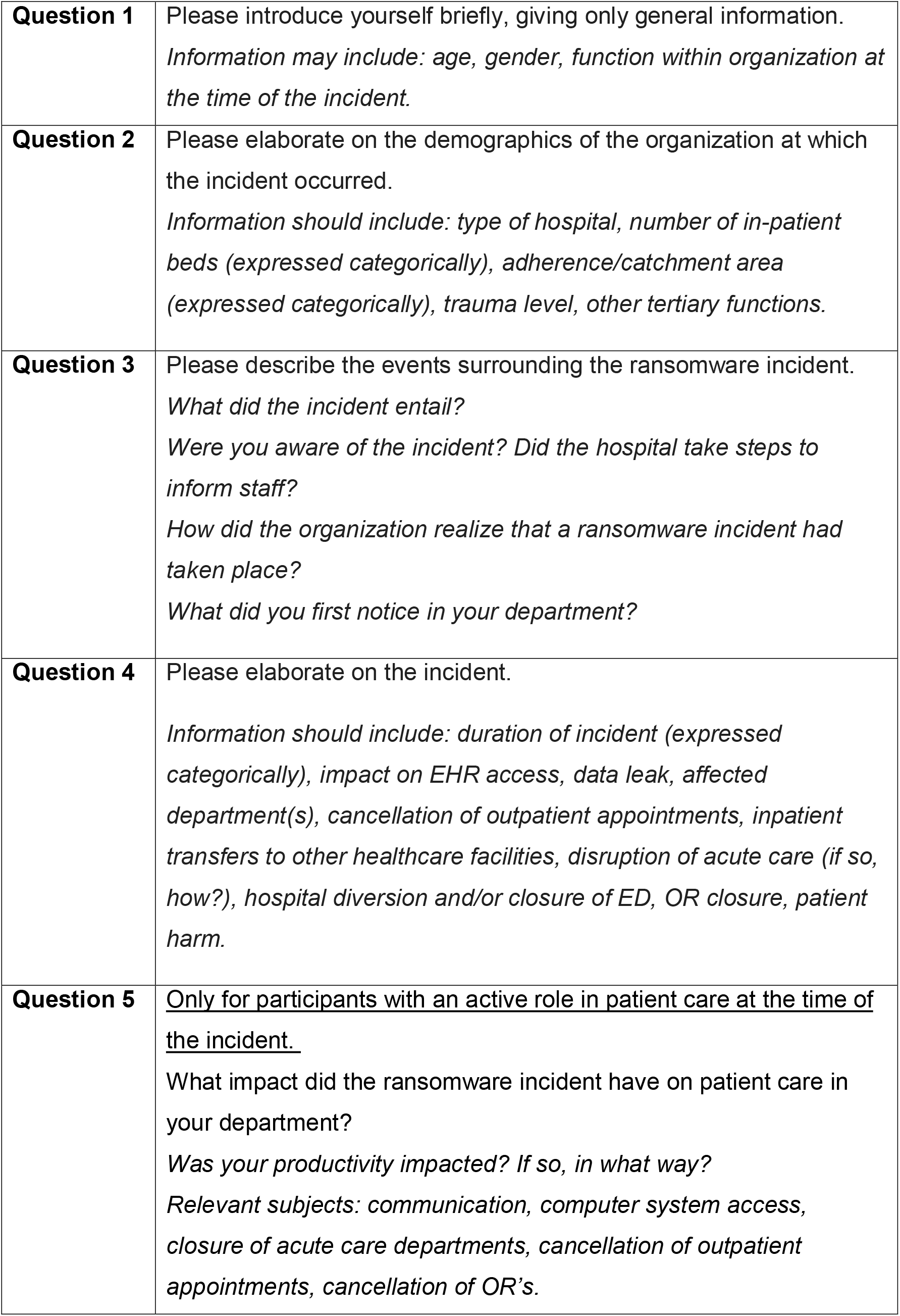
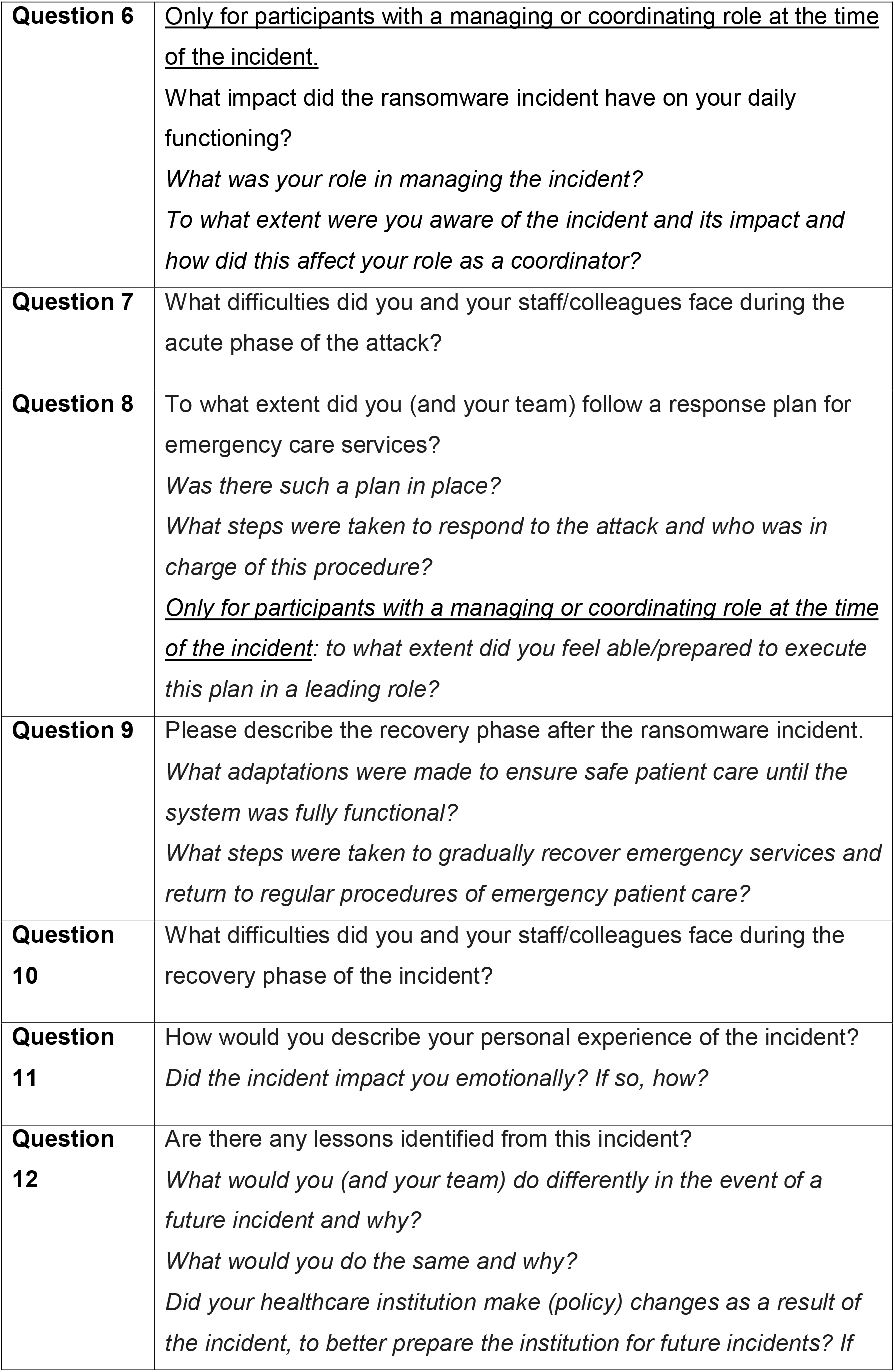
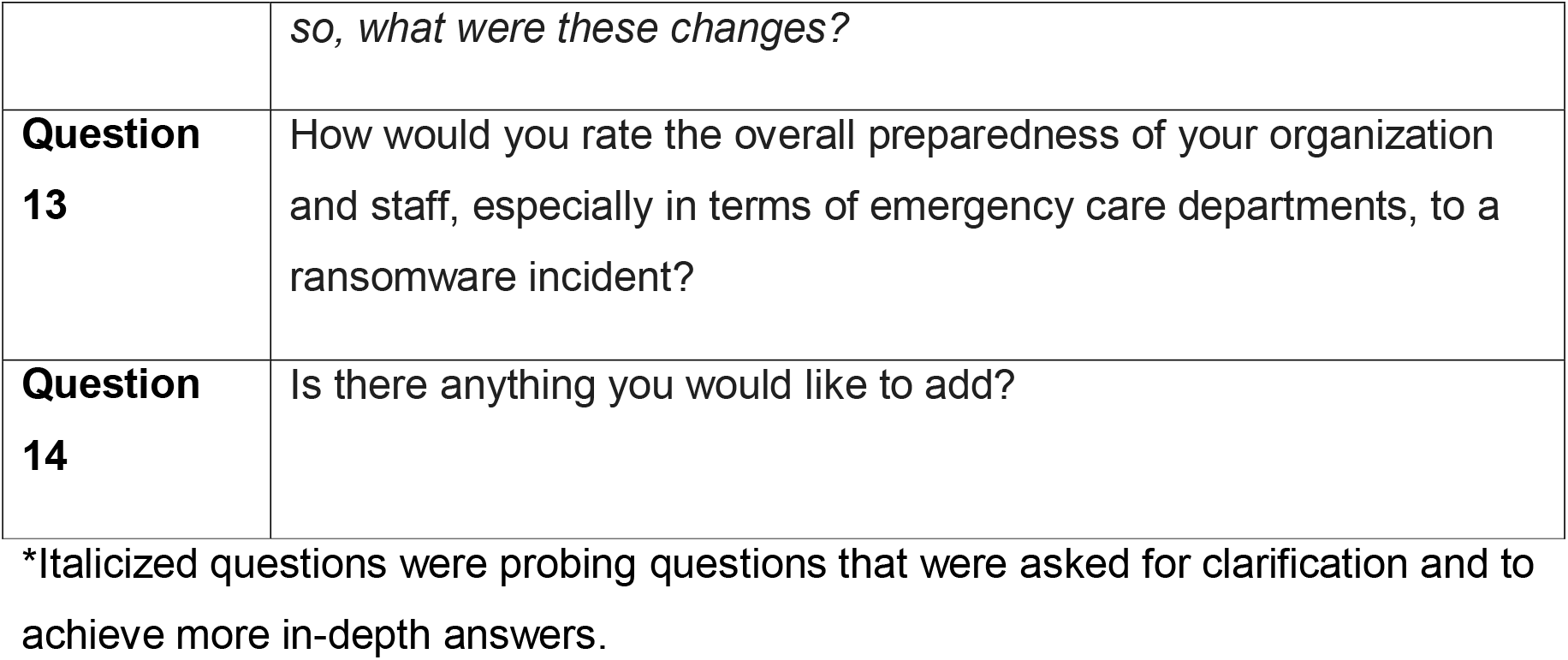
Interview Guideline

|  |  |
| --- | --- |
| **Question 1** | Please introduce yourself briefly, giving only general information.<br>*Information may include: age, gender, function within organization at the time of the incident.* |
| **Question 2** | Please elaborate on the demographics of the organization at which the incident occurred.<br>*Information should include: type of hospital, number of in-patient beds (expressed categorically), adherence/catchment area (expressed categorically), trauma level, other tertiary functions.* |
| **Question 3** | Please describe the events surrounding the ransomware incident.<br>*What did the incident entail?*<br>*Were you aware of the incident? Did the hospital take steps to inform staff?*<br>*How did the organization realize that a ransomware incident had taken place?*<br>*What did you first notice in your department?* |
| **Question 4** | Please elaborate on the incident.<br>*Information should include: duration of incident (expressed categorically), impact on EHR access, data leak, affected department(s), cancellation of outpatient appointments, inpatient transfers to other healthcare facilities, disruption of acute care (if so, how?), hospital diversion and/or closure of ED, OR closure, patient harm.* |
| **Question 5** | <u>Only for participants with an active role in patient care at the time of the incident.</u><br>What impact did the ransomware incident have on patient care in your department?<br>*Was your productivity impacted? If so, in what way?*<br>*Relevant subjects: communication, computer system access, closure of acute care departments, cancellation of outpatient appointments, cancellation of OR's.* |
| **Question 6** | <u>Only for participants with a managing or coordinating role at the time of the incident.</u><br>What impact did the ransomware incident have on your daily functioning?<br>*What was your role in managing the incident?*<br>*To what extent were you aware of the incident and its impact and how did this affect your role as a coordinator?* |
| **Question 7** | What difficulties did you and your staff/colleagues face during the acute phase of the attack? |
| **Question 8** | To what extent did you (and your team) follow a response plan for emergency care services?<br>*Was there such a plan in place?*<br>*What steps were taken to respond to the attack and who was in charge of this procedure?*<br><u>Only for participants with a managing or coordinating role at the time of the incident:</u> *to what extent did you feel able/prepared to execute this plan in a leading role?* |
| **Question 9** | Please describe the recovery phase after the ransomware incident.<br>*What adaptations were made to ensure safe patient care until the system was fully functional?*<br>*What steps were taken to gradually recover emergency services and return to regular procedures of emergency patient care?* |
| **Question 10** | What difficulties did you and your staff/colleagues face during the recovery phase of the incident? |
| **Question 11** | How would you describe your personal experience of the incident?<br>*Did the incident impact you emotionally? If so, how?* |
| **Question 12** | Are there any lessons identified from this incident?<br>*What would you (and your team) do differently in the event of a future incident and why?*<br>*What would you do the same and why?*<br>*Did your healthcare institution make (policy) changes as a result of the incident, to better prepare the institution for future incidents? If* |
|  | *so, what were these changes?* |
| **Question**<br>**13** | How would you rate the overall preparedness of your organization and staff, especially in terms of emergency care departments, to a ransomware incident? |
| **Question**<br>**14** | Is there anything you would like to add? |

\*Italicized questions were probing questions that were asked for clarification and to achieve more in-depth answers.

